# PATTERNS OF OCCUPATIONAL MORBIDITY IN ZAMBIA, 2008-2018: A DESCRIPTIVE DATABASE STUDY

**DOI:** 10.1101/2021.04.27.21255681

**Authors:** M. Zambwe, P.C. Bwembya, R. Mutemwa, J. Gasana

## Abstract

**OBJECTIVE:** This study aimed to describe characteristics of occupational morbidity in Zambia over an eleven year period: 2008-2018.

**METHODS:** A descriptive retrospective observational study based on compensation claims database from the Workers Compensation Fund Control Board (WCFCB) in Zambia over the period 2008 to 2018 was conducted. All the accepted compensation claims at WCFCB during the period 2008 to 2018 were reviewed. The reference population of the study was all workers in Zambia covered by WCFCB at the time. All the accepted compensation claims during the period were taken up into the study. Stata version 14 was used to analyze the data, and make descriptive tables and graphs.

**RESULTS:** The total number of reviewed and analyzed cases was 8,009. The gender most affected by occupational morbidity was males (94%). Married males were more affected (72%) compared to single males. However, single females were more affected than the married females at 4% and 2%, respectively. The major morbidity types were wounds (30%), fractures (29%), and amputations (17%). The biggest contributing industries to the cause of occupational morbidity were manufacturing (27%), and mining (19%). Lusaka and Copperbelt regions were the main epicenters at 49% and 34% respectively.

**CONCLUSION:** Wounds, fractures and amputations were the most prominent types of occupational morbidity. Traditional gender-based practices of married males seem to underlie their over-exposure to occupational hazards. Manufacturing sector which is poorly regulated compared to the mining sector, was the highest contributor to the occupational morbidity. The manufacturing sector should be subjected to stronger government regulation and inspectorate, with emphasis on compliance to relevant international occupational health and safety protocols.

**ARTICLE SUMMARY:** *Strengths and limitations of the study:*

- The study utilized the WCFCB injury and disease compensation claims data as a proxy to establish the national burden of occupational morbidity in Zambia during the study period, hence contributing to the body of knowledge.
- WCFCB does not cover government ministries and the informal sector in the country which accounts for about 89.3% of the total labour force, the injuries and diseases being contracted in the foretasted sectors were never reported to the institution; and hence making findings of our study an understatement of the actual national burden.
- Since submission of compensation claims is generally motivated by receiving monetary benefits for an injury or disease contracted, cases of less magnitude are not reported to WCFCB, hence could not be captured in the study.
- Out of many diseases WCFCB only recognize pneumoconiosis and pulmonary tuberculosis as occupational diseases, hence our study could only assess those two making the study an understatement of the actual occupational disease burden.

## INTRODUCTION

Occupational injuries are one of the top ten leading causes of morbidity and mortality globally.^1^ Each year about 313 million workers around the world get involved in occupational non-fatal accidents; while about 160 million workers contract non-fatal occupational diseases.^2^ And each year about 2.3 million workers in the world die from occupational injuries and diseases. Specifically about 350,000 workers die from injuries; and about 2 million die from occupational diseases each year globally.^2^

The distribution of the 2014 global fatal and non-fatal occupational accidents over the five WHO regions was that: Asia had the highest number of fatalities constituting more than 70% of the global share with an occupational fatality rate of 12.7 per 100,000 persons. Africa was second rating; though it had the highest occupational fatality rate of 16.6 per 100,000 persons.^3^

In their study of mortality burden in Zambia, CSO (2018) found that out of the top ten leading causes of death in in all age groups in Zambia, accidents and injuries were the fourth at 10%.^4^ Within the employable age group of 15 years and above, the same study found that injuries occupied the third slot at 11%.

The causes of occupational injuries are diverse and complicated. But research has identified some of them as: manually delivered jobs and longer working hours.^5^ Researchers have also found associations of injuries with race, age, gender, education and geographical location.^6^ Also younger workers have most been found to lack work experience which increases their likelihood of getting occupational injuries.^7^

From the fore-going it is evident that occupational injuries and diseases are surely a major public health challenge globally. They can end up in disability, loss of employment, homelessness and even mortality in some cases.^8^ Therefore, occupational injuries deserve a huge recognition that goes with effective and efficient interventions to save millions of lives being lost each day that passes.

To the best of knowledge there is a general lack of studies that profile the national burden of occupational morbidity in Zambia. Most studies seen would only focus on a single particular occupation, morbidity type, or location; and of shorter periods. This study was therefore undertaken to help quantify the magnitude of this public health challenge in Zambia by covering all its major characteristics and over an extended period of time. The study will therefore contribute to raising awareness and making the public health challenge of occupational injuries and diseases in Zambia more visible to managers and policy makers in private companies, public companies, non-governmental organizations and the government. The increased visibility of the problem will in turn help the managers and policy makers formulate effective and efficient intervention strategies that will save thousands of lives in the country that are each year being lost to disabilities and death.

Determining the national burden of occupational injuries and diseases in any country can often be problematic. This is so because most countries around the world, Zambia inclusive, do not have national reporting centers for all occupational morbidities and mortalities. This being the case, the practice in most countries around the world is to use the Workers Compensation claims data as a proxy.^9^

The study focused on all workers in Zambia who filed in accepted compensation claims with the Workers Compensation Fund Control Board (WCFCB) between the years 2008 and 2018. Such workers must have been injured, contracted a disease or died while participating in an economically productive activity for wages or profit within the above stated period.

The objective of this study was therefore to establish the patterns of occupational morbidity in Zambia over the period 2008 to 2018 by sex, marital status, age morbidity type, industry and location. WCFCB claims data were used.

## METHODS

A descriptive retrospective observational database study design was applied. Accepted occupational injury and disease compensation claim records which were submitted to WCFCB over the period January 1 2008 to December 31 2018 were examined. A data extraction tool was used to record relevant data from the claimant records.

The study was conducted on WCFCB which operates twenty branches over the ten provinces of the country. So the study setting was the ten provinces of the republic of Zambia within the period 2008 - 2018. WCFCB is a government institution under the Ministry of Labour. Its mandate is to pool funds from employers, for the purpose of financing occupational healthcare of injured and diseased workers, and pay them compensations for the resultant loss of physiological function or death.

WCFCB operates an Oracle based internet propelled digital system to register and process compensation claims. The system is called the Pension Administration System (PAS) version 1.5. All the branches, regional offices and head office of WCFCB are connected to the PAS system. And at every branch is a Benefits Officer whose duty is to receive compensation claims and enroll them into the system which subsequently processes them into relevant information used to pay compensations and finance occupational healthcare of the claimants.

This study was approved by the Ethics Committee of the WCFCB. Upon formal request to WCFCB management, a summary report of all accepted claims in the period was issued. And a data extraction tool was used to collect relevant information from the summary report. Further epidemiological details of the cases were obtained by accessing their individual digital records on PAS. With the approval of the WCFCB management we extracted data from their PAS system with the following attributes sex, marital status, age, morbidity type, industry and region.

### Inclusion and exclusion criteria

All the occupational injury and disease claim cases that were submitted and accepted by WCBCB over the period 2008 to 2018 were extracted, reviewed and included in the study. The total number of all such cases was 8,009. We analyzed sex, marital status, age, morbidity type, industry and region. The age was split into the following age groups 25-24, 25-35, 36-49, 50-59, and ≥60. The industries from which the compensation claims were reported were classified according to the United Nations International Standard Industrial Classification of All Economic Activities (ISIC). And eight groups namely mining, transport, agriculture, construction, energy, manufacturing, support services and trade were identified.

The study excluded the following:

- subjects who did not file in occupational injury and disease compensation claims with WCFCB within the study period.
- subjects whose compensation claims were not accepted by WCFCB within the study period.
- cases from the informal sector, government ministries, government defense and security sectors

### Statistical analysis

Data from the data extraction tool were entered into a Microsoft excel sheet, checked for duplication and subsequently exported to Stata version 14 for data analysis using descriptive statistics alone. One frequency table with actual variable counts and their corresponding percentages was created. Five bar graphs with percent frequencies were further created.

### Patient and public involvement

We did not involve patients and public in the study. We simply extracted their anonymised data from the WCFCB database.

### Definition of Terms

|  |  |
| --- | --- |
| Worker | Any person who works for another from whom he receives remuneration.<sup>10</sup> |
| Employer | Any person who provides work to another and remunerates them.<sup>10</sup> |
| Occupational Disease | A disease contracted as a result of one's exposure to occupational risk factors.<sup>11</sup> In this study it only refers to pneumoconiosis and pulmonary tuberculosis which are both lung diseases. |
| Pension Administration System (PAS) | The Workers Compensation Fund control Board system that enrolls and processes occupational injury and disease Compensation claims. It is Oracle based and internet propelled. |
| Workers Compensation Fund Control Board | The mandatory statutory insurance scheme for all workers in Zambia created by the Zambian Parliament under the Workers Compensation Act 10 of 1999. Its mandate is to finance the healthcare of all occupational injuries and diseases of workers and compensate them for the resultant loss of physiological function or death. |

## RESULTS

There were a total of 8 009 occupational morbidity cases reported and accepted by WCFCB as compensation claim cases over the eleven year period 2008 to 2018. The majority of the cases occurred among the males (7 528; 94%), and among the marrieds (5 927; 74%), in the age group 25-35 years (3 260; 41%). The most common morbidity type during the period was wounds (2 390; 30%), mostly affecting the manufacturing industry (2 170; 27%). And Lusaka province (3 935; 49%) was the most affected among all the regions in the country.

In Table 1, the sex most affected by occupational injuries and diseases was found to be males with a frequency of 94% over the study period. The marital status most affected were the married at 74%. And the age group most affected by occupational morbidity was the 25-35 years.

**Table 1:**
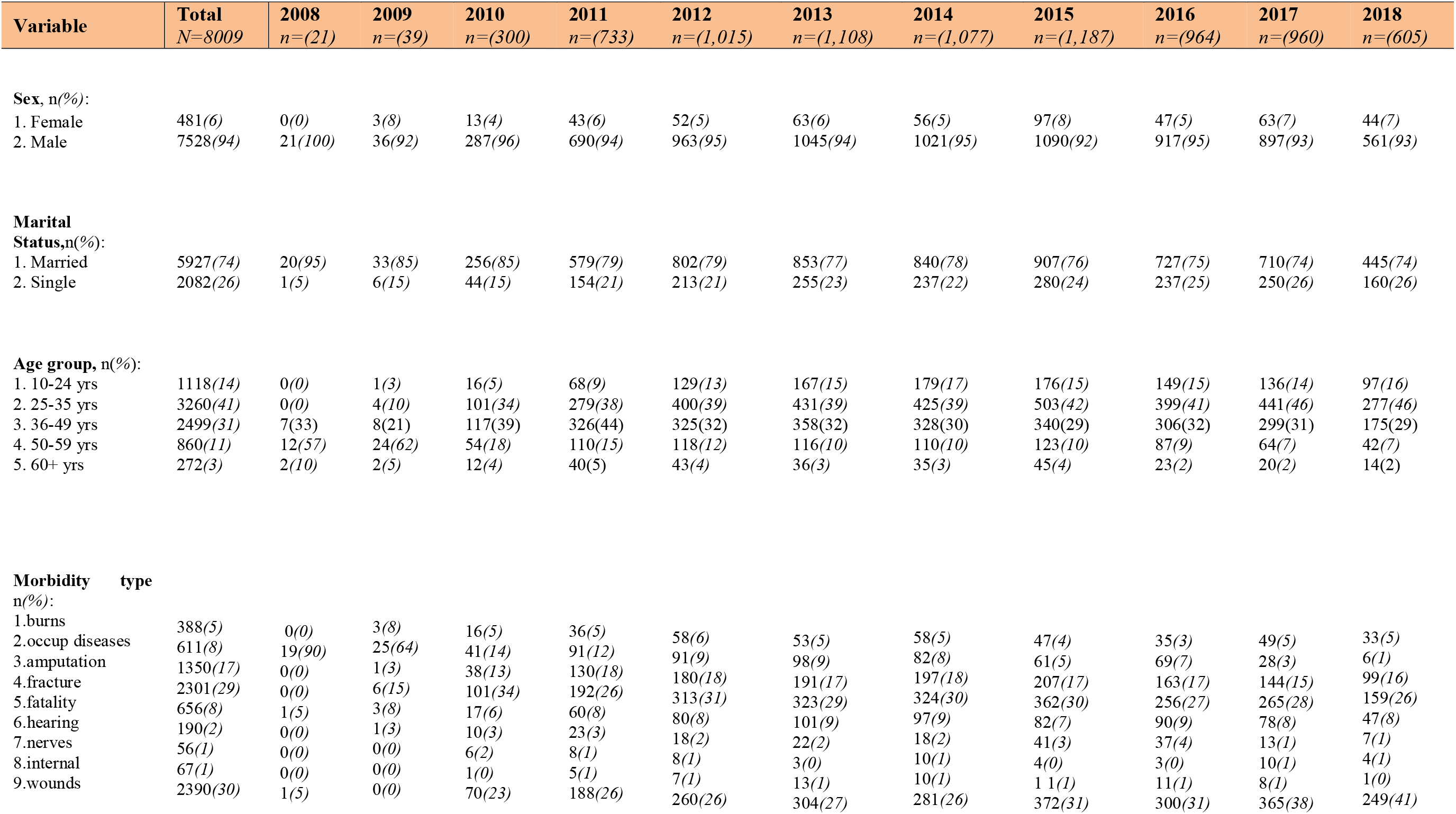
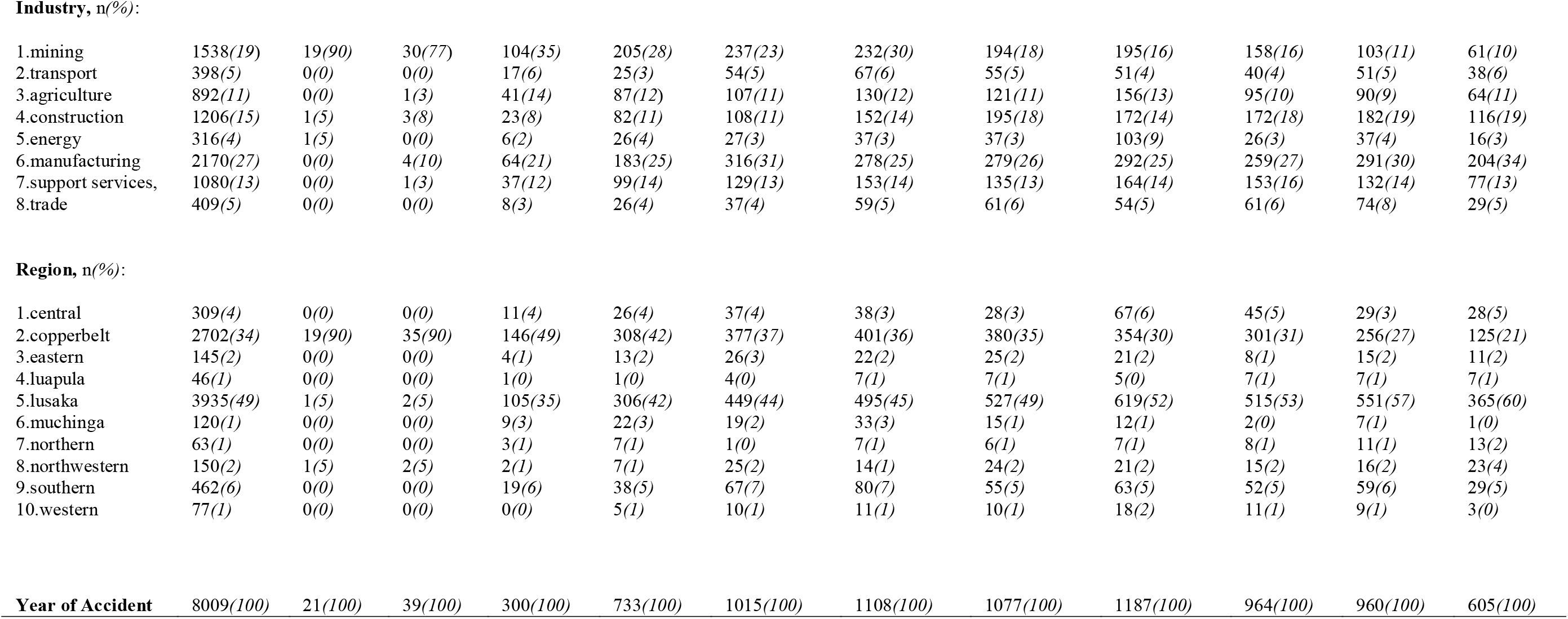
Profile of occupational morbidities in Zambian over the period 2008 – 2018.

| Variable | Total<br>N=8009 | 2008<br>n=(21) | 2009<br>n=(39) | 2010<br>n=(300) | 2011<br>n=(733) | 2012<br>n=(1,015) | 2013<br>n=(1,108) | 2014<br>n=(1,077) | 2015<br>n=(1,187) | 2016<br>n=(964) | 2017<br>n=(960) | 2018<br>n=(605) |
| --- | --- | --- | --- | --- | --- | --- | --- | --- | --- | --- | --- | --- |
| **Sex, n(%):** |  |  |  |  |  |  |  |  |  |  |  |  |
| 1. Female | 481(6) | 0(0) | 3(8) | 13(4) | 43(6) | 52(5) | 63(6) | 56(5) | 97(8) | 47(5) | 63(7) | 44(7) |
| 2. Male | 7528(94) | 21(100) | 36(92) | 287(96) | 690(94) | 963(95) | 1045(94) | 1021(95) | 1090(92) | 917(95) | 897(93) | 561(93) |
| **Marital Status,n(%):** |  |  |  |  |  |  |  |  |  |  |  |  |
| 1. Married | 5927(74) | 20(95) | 33(85) | 256(85) | 579(79) | 802(79) | 853(77) | 840(78) | 907(76) | 727(75) | 710(74) | 445(74) |
| 2. Single | 2082(26) | 1(5) | 6(15) | 44(15) | 154(21) | 213(21) | 255(23) | 237(22) | 280(24) | 237(25) | 250(26) | 160(26) |
| **Age group, n(%):** |  |  |  |  |  |  |  |  |  |  |  |  |
| 1. 10-24 yrs | 1118(14) | 0(0) | 1(3) | 16(5) | 68(9) | 129(13) | 167(15) | 179(17) | 176(15) | 149(15) | 136(14) | 97(16) |
| 2. 25-35 yrs | 3260(41) | 0(0) | 4(10) | 101(34) | 279(38) | 400(39) | 431(39) | 425(39) | 503(42) | 399(41) | 441(46) | 277(46) |
| 3. 36-49 yrs | 2499(31) | 7(33) | 8(21) | 117(39) | 326(44) | 325(32) | 358(32) | 328(30) | 340(29) | 306(32) | 299(31) | 175(29) |
| 4. 50-59 yrs | 860(11) | 12(57) | 24(62) | 54(18) | 110(15) | 118(12) | 116(10) | 110(10) | 123(10) | 87(9) | 64(7) | 42(7) |
| 5. 60+ yrs | 272(3) | 2(10) | 2(5) | 12(4) | 40(5) | 43(4) | 36(3) | 35(3) | 45(4) | 23(2) | 20(2) | 14(2) |
| **Morbidity type n(%):** |  |  |  |  |  |  |  |  |  |  |  |  |
| 1.burns | 388(5) | 0(0) | 3(8) | 16(5) | 36(5) | 58(6) | 53(5) | 58(5) | 47(4) | 35(3) | 49(5) | 33(5) |
| 2.occup diseases | 611(8) | 19(90) | 25(64) | 41(14) | 91(12) | 91(9) | 98(9) | 82(8) | 61(5) | 69(7) | 28(3) | 6(1) |
| 3.amputation | 1350(17) | 0(0) | 1(3) | 38(13) | 130(18) | 180(18) | 191(17) | 197(18) | 207(17) | 163(17) | 144(15) | 99(16) |
| 4.fracture | 2301(29) | 0(0) | 6(15) | 101(34) | 192(26) | 313(31) | 323(29) | 324(30) | 362(30) | 256(27) | 265(28) | 159(26) |
| 5.fatality | 656(8) | 1(5) | 3(8) | 17(6) | 60(8) | 80(8) | 101(9) | 97(9) | 82(7) | 90(9) | 78(8) | 47(8) |
| 6.hearing | 190(2) | 0(0) | 1(3) | 10(3) | 23(3) | 18(2) | 22(2) | 18(2) | 41(3) | 37(4) | 13(1) | 7(1) |
| 7.nerves | 56(1) | 0(0) | 0(0) | 6(2) | 8(1) | 8(1) | 3(0) | 10(1) | 4(0) | 3(0) | 10(1) | 4(1) |
| 8.internal | 67(1) | 0(0) | 0(0) | 1(0) | 5(1) | 7(1) | 13(1) | 10(1) | 11(1) | 11(1) | 8(1) | 1(0) |
| 9.wounds | 2390(30) | 1(5) | 0(0) | 70(23) | 188(26) | 260(26) | 304(27) | 281(26) | 372(31) | 300(31) | 365(38) | 249(41) |
| **Industry, n(%):** |  |  |  |  |  |  |  |  |  |  |  |  |
| 1.mining | 1538(19) | 19(90) | 30(77) | 104(35) | 205(28) | 237(23) | 232(30) | 194(18) | 195(16) | 158(16) | 103(11) | 61(10) |
| 2.transport | 398(5) | 0(0) | 0(0) | 17(6) | 25(3) | 54(5) | 67(6) | 55(5) | 51(4) | 40(4) | 51(5) | 38(6) |
| 3.agriculture | 892(11) | 0(0) | 1(3) | 41(14) | 87(12) | 107(11) | 130(12) | 121(11) | 156(13) | 95(10) | 90(9) | 64(11) |
| 4.construction | 1206(15) | 1(5) | 3(8) | 23(8) | 82(11) | 108(11) | 152(14) | 195(18) | 172(14) | 172(18) | 182(19) | 116(19) |
| 5.energy | 316(4) | 1(5) | 0(0) | 6(2) | 26(4) | 27(3) | 37(3) | 37(3) | 103(9) | 26(3) | 37(4) | 16(3) |
| 6.manufacturing | 2170(27) | 0(0) | 4(10) | 64(21) | 183(25) | 316(31) | 278(25) | 279(26) | 292(25) | 259(27) | 291(30) | 204(34) |
| 7.support services, | 1080(13) | 0(0) | 1(3) | 37(12) | 99(14) | 129(13) | 153(14) | 135(13) | 164(14) | 153(16) | 132(14) | 77(13) |
| 8.trade | 409(5) | 0(0) | 0(0) | 8(3) | 26(4) | 37(4) | 59(5) | 61(6) | 54(5) | 61(6) | 74(8) | 29(5) |
| **Region, n(%):** |  |  |  |  |  |  |  |  |  |  |  |  |
| 1.central | 309(4) | 0(0) | 0(0) | 11(4) | 26(4) | 37(4) | 38(3) | 28(3) | 67(6) | 45(5) | 29(3) | 28(5) |
| 2.copperbelt | 2702(34) | 19(90) | 35(90) | 146(49) | 308(42) | 377(37) | 401(36) | 380(35) | 354(30) | 301(31) | 256(27) | 125(21) |
| 3.eastern | 145(2) | 0(0) | 0(0) | 4(1) | 13(2) | 26(3) | 22(2) | 25(2) | 21(2) | 8(1) | 15(2) | 11(2) |
| 4.luapula | 46(1) | 0(0) | 0(0) | 1(0) | 1(0) | 4(0) | 7(1) | 7(1) | 5(0) | 7(1) | 7(1) | 7(1) |
| 5.lusaka | 3935(49) | 1(5) | 2(5) | 105(35) | 306(42) | 449(44) | 495(45) | 527(49) | 619(52) | 515(53) | 551(57) | 365(60) |
| 6.muchinga | 120(1) | 0(0) | 0(0) | 9(3) | 22(3) | 19(2) | 33(3) | 15(1) | 12(1) | 2(0) | 7(1) | 1(0) |
| 7.northern | 63(1) | 0(0) | 0(0) | 3(1) | 7(1) | 1(0) | 7(1) | 6(1) | 7(1) | 8(1) | 11(1) | 13(2) |
| 8.northwestern | 150(2) | 1(5) | 2(5) | 2(1) | 7(1) | 25(2) | 14(1) | 24(2) | 21(2) | 15(2) | 16(2) | 23(4) |
| 9.southern | 462(6) | 0(0) | 0(0) | 19(6) | 38(5) | 67(7) | 80(7) | 55(5) | 63(5) | 52(5) | 59(6) | 29(5) |
| 10.western | 77(1) | 0(0) | 0(0) | 0(0) | 5(1) | 10(1) | 11(1) | 10(1) | 18(2) | 11(1) | 9(1) | 3(0) |
| **Year of Accident** | 8009(100) | 21(100) | 39(100) | 300(100) | 733(100) | 1015(100) | 1108(100) | 1077(100) | 1187(100) | 964(100) | 960(100) | 605(100) |

### Sex and marital status

In Figure 1, males were the most affected by occupational morbidities at a total sum of 94.3%. The married males were most affected over the single males at 71.6% and 22.7% respectively. The percent of females affected was only 5.7%. Whereas the marrieds were the most affected among the males; the singles were the most affected at 3.6% among the female workers.

**Fig 1:**
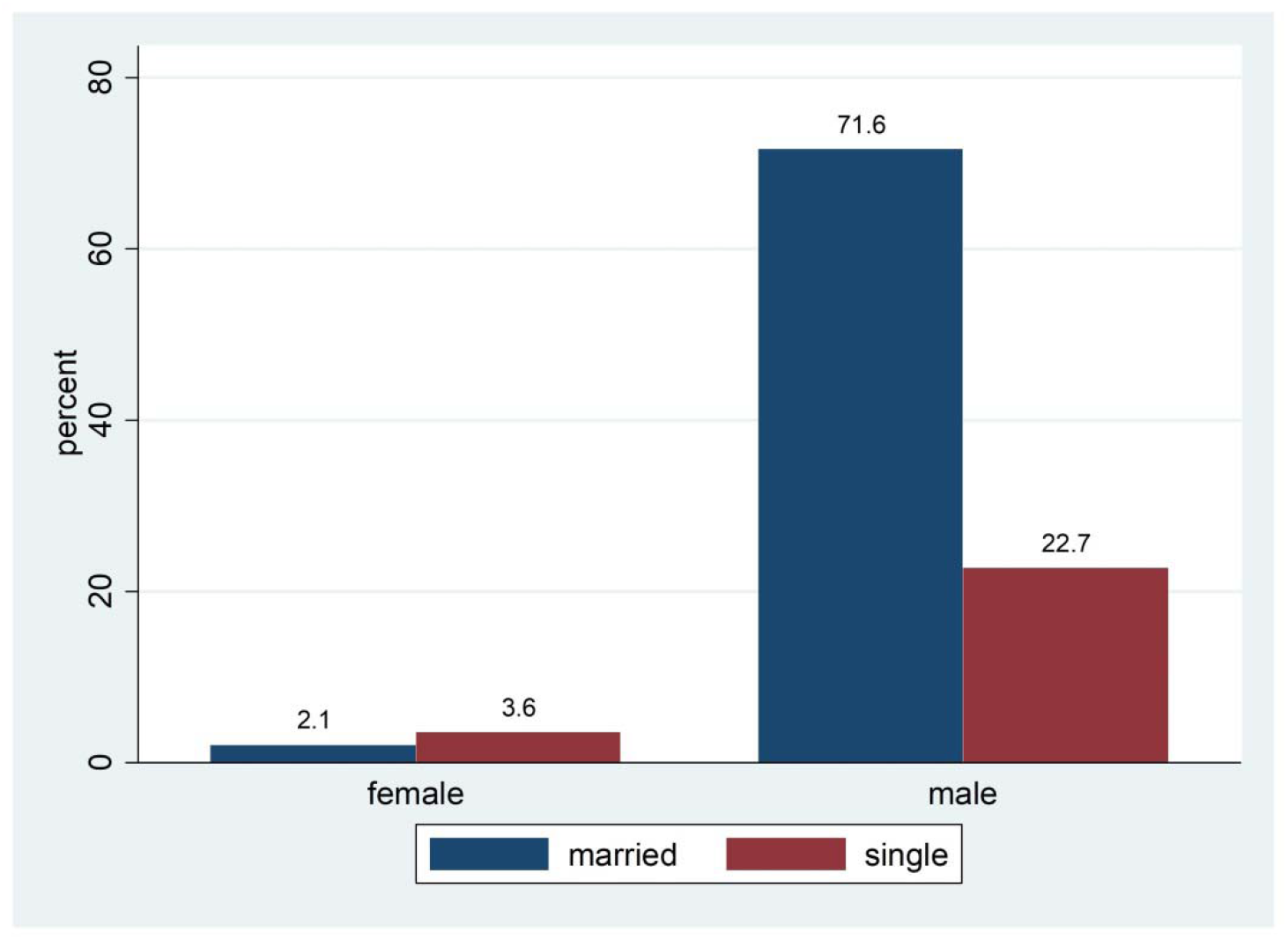
Occupational morbidities by sex and marital status in Zambia over the period 2008 to 2018.

### Age group and summary outcome

In Figure 2, occupational diseases affected the 36 to 49 years age group, the most at 3.3%. They appeared slow setting reaching their peak impact in the age group 36 to 49 years. Thereafter they continued to decline with age. The 50 to 59 years age group was the second most affected at 2.5%. At above 60 years the cases of occupational diseases among workers were almost nil.

**Fig 2:**
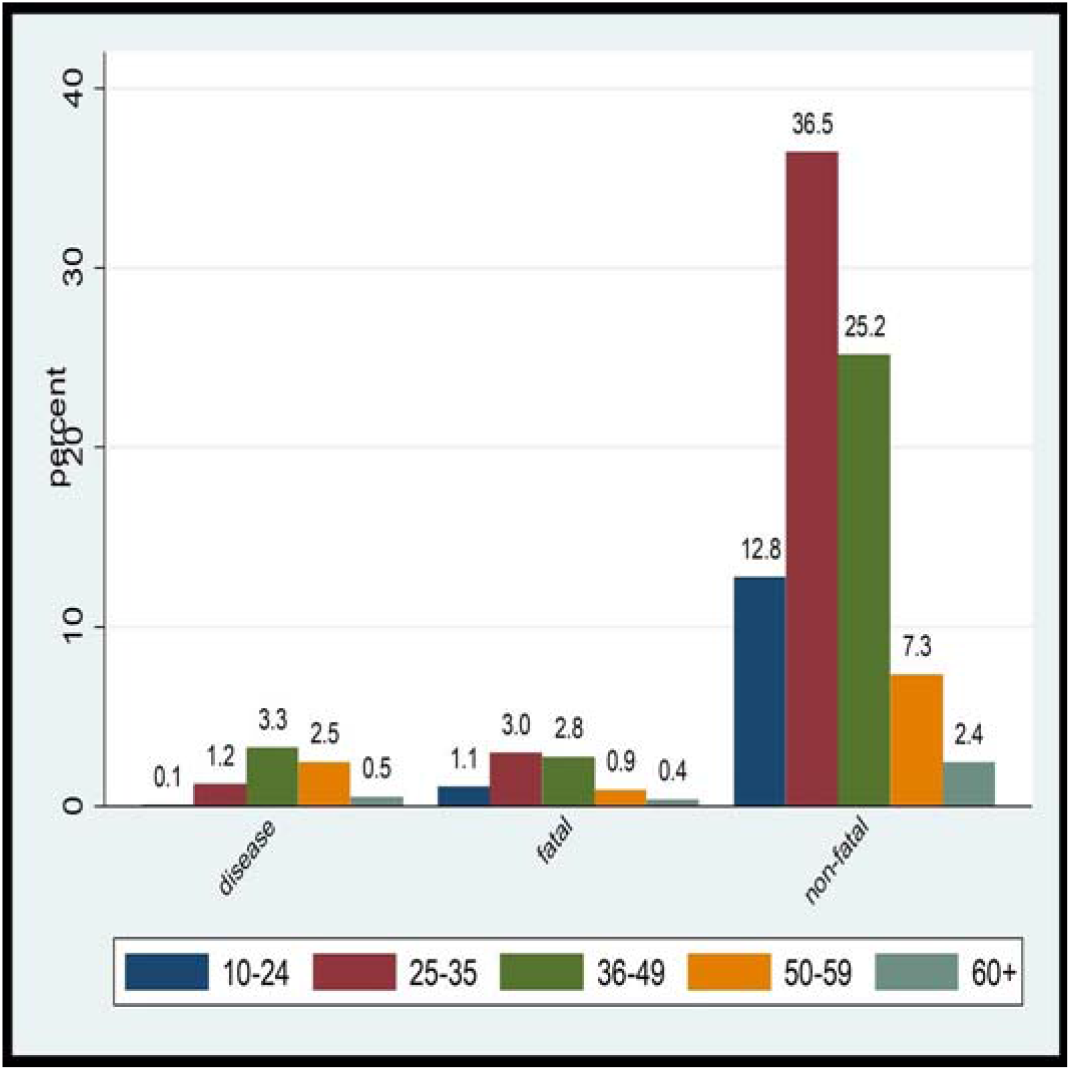
Occupational morbidities by summary outcome by age group in Zambia over the period 2008 to 2018.

Fatal injuries and non-fatal injuries were found to be fast setting with age. The frequency increased with age of workers. They were found to reach their peak impact in age group 25 to 35 years at 39.5%. Thereafter they continued to decline with age to about 2.4% for workers above 60 years of age.

### Occupational morbidity types

In Figure 3, wounds were the most common type of occupational morbidity suffered by workers at about 30% in the period, closely followed by fractures at about 29%, and amputations at about 17%.

**Figure 3:**
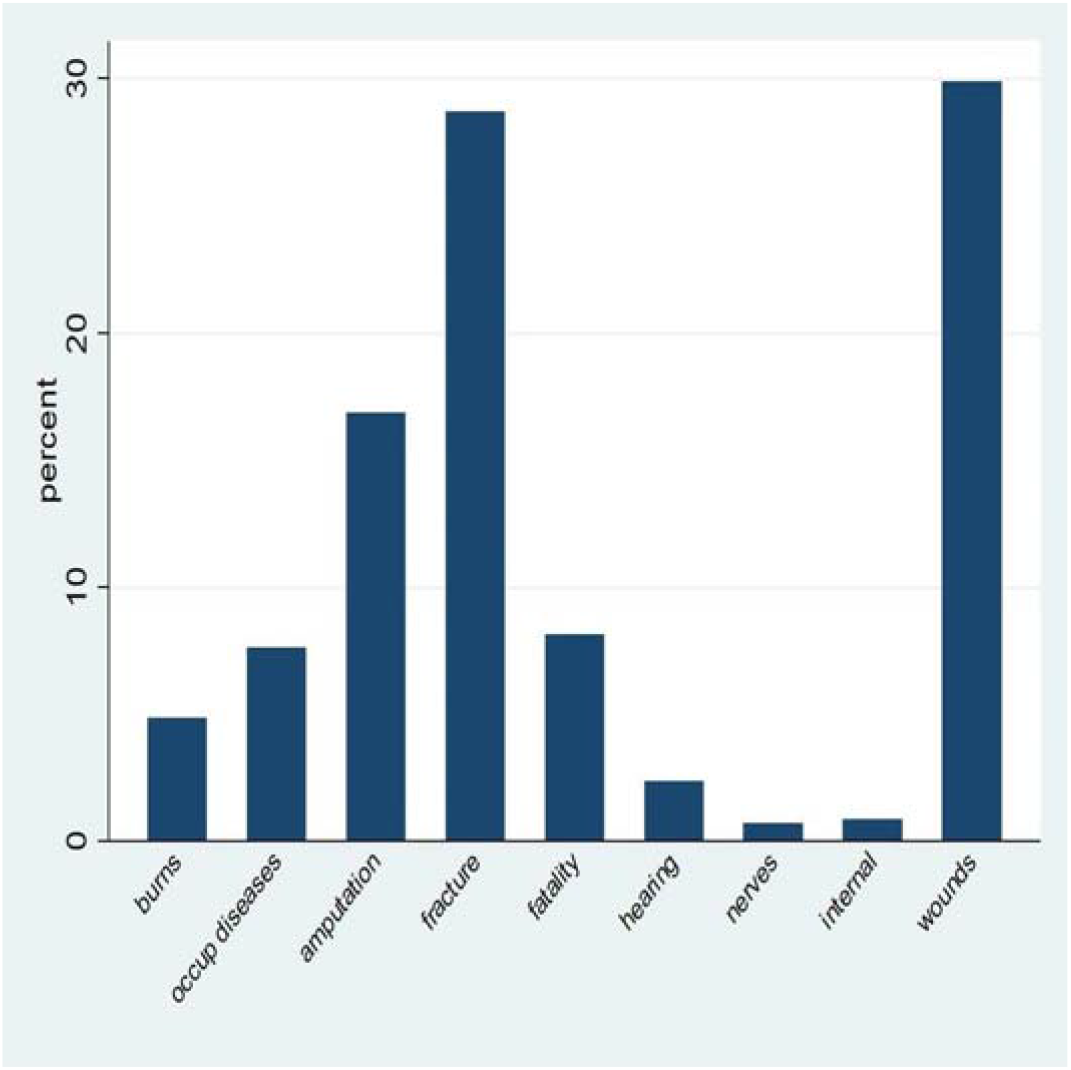
Occupational morbidity by morbidity types in Zambia over the period 2008 to 2018.

### Occupational morbidity by industrial sector

In Figure 4, the morbidity burden by industrial sector indicated manufacturing as the leading contributor at about 26%. The mining industry was second with about 19% cases over the period. Construction and support services were the third and fourth at about 15% and 13% respectively.

**Figure 4:**
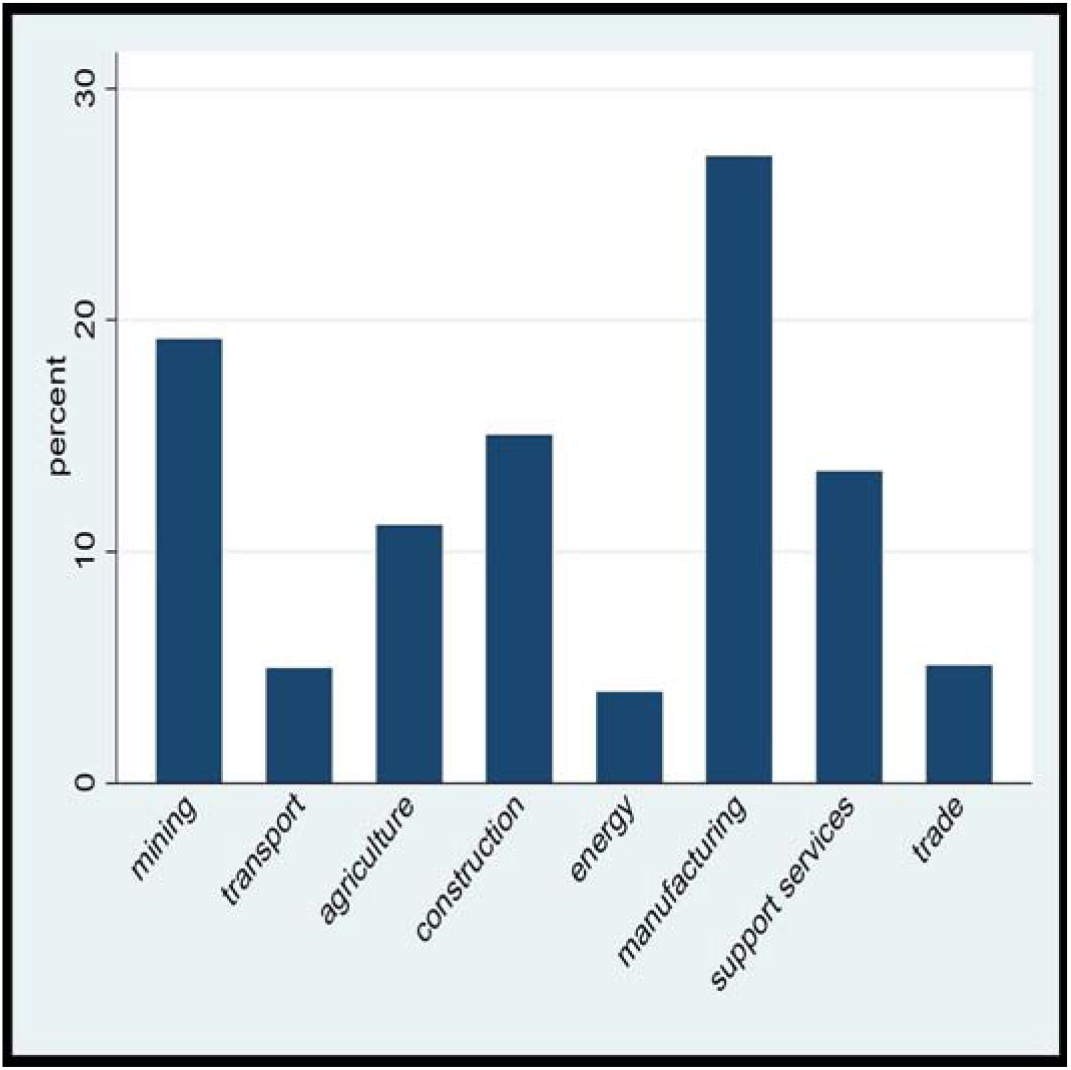
Occupational morbidity by industrial sector in Zambia over the period 2008 to 2018.

### Occupational morbidity by region

In Figure 5, among all the regions, Lusaka was the most affected with 49.1% incidence of the cases. Copperbelt was the second most impacted with 33.7%. Southern region took the third place at 5.8%; and Luapula was the least affected with only 0.6% cases over the period.

**Figure 5:**
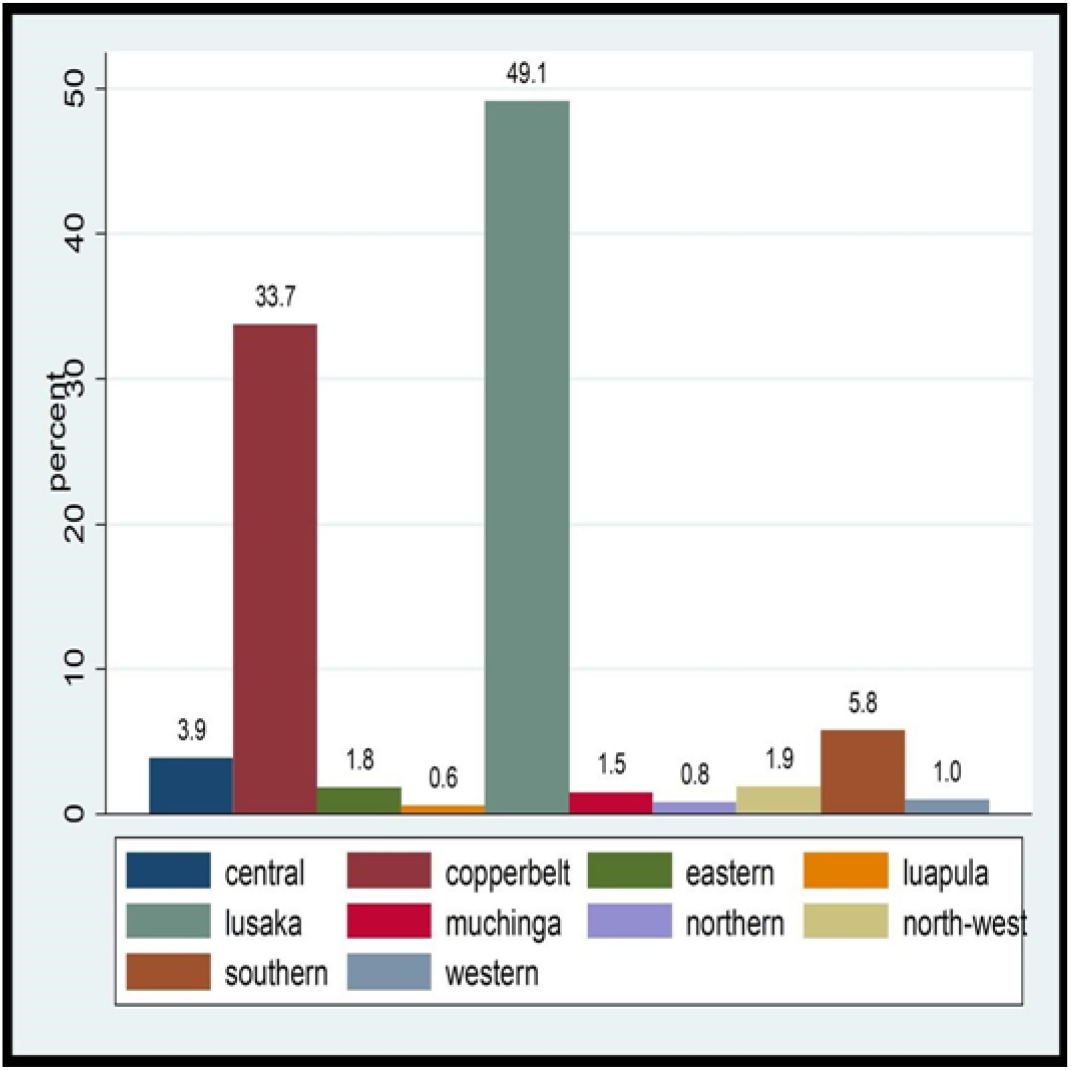
Occupational morbidities by region over the period 2008 to 2018.

## DISCUSSION

Out of all the studies of occupational morbidity conducted in Zambia, this study covered the whole country with the highest number of cases over the longest period of time. Our study based on the occupational compensation claims database of the WCFCB set out to establish patterns of occupational morbidity in Zambia. The focus of the study was a full scope of the challenge of occupational morbidities in the country by sex, marital status, age of workers, morbidity type, industry, and location of the workers. The mining industry is traditionally deemed the most hazardous of all sectors. However, in our study manufacturing was more prominent and more hazardous than any other sectors in the country.

### Sex, marital status and age group

With 94% claim rate coming from the males; they were the most affected and most vulnerable gender to occupational morbidities. This was consistent with similar studies conducted in India, South Australia, Kitwe-Zambia, and Lusaka-Zambia.^12–15^

Our study further observed that marital status was one of the key factors to the incidence of occupational injuries and diseases. We established that 78% of all the cases were married. This result concurred with similar studies in Australia, Iran and Southern Ethiopia.^13,16 17^ Matching marital status with sex, our study found that the majority cases were found to be married males. The reason for this phenomenon was unclear but we may argue that socio-economic patterns imposed by economic hardship at household level could be the most responsible factor. For instance, single females were found to be more affected than their married female counterparts. The reason is very likely that traditional gender practices have tended to render it easier for the single woman (even with a child) to exercise her right to get a job to support herself, compared to the married woman who has a man in her life to “look after her”. Some men actively forbid their wives from seeking employment, especially when the wife is still young and having children. To some degree, this result mimics similar findings from a study in the United Kingdom on accidents risks, gender, family status and occupation choices; where they established that workers with children are more risk averse and more likely to avoid risky jobs than workers without.^18^ Going by the above argument, the married females in our study were more likely to have children and hence their more risk aversion resulting in lesser accident cases than the single females who might have had no children and hence their less risk aversion presenting itself in higher incidence of injuries and diseases.

Overall the most affected age grouping in the study was the 25 to 35 years with a 41% incidence rate. And going by the specific outcomes, fatal injuries and non-fatal injuries were most rampant in the age group above at 36.6% and 43.3% respectively. These results were very much in agreement with other studies on the occupational safety profile in Zambia, compensation patterns in Zambia, in Southern China, Uganda, and Ghana.^10,19–22^ However, for occupational diseases our study established that the most vulnerable age group was the 36 to 49 years at 42.9%. This was consistent with study findings among underground gold miners in Ghana; and among open pit copper miners in Chingola.^23 24^ However, two studies by Sitembo (2012) and Leigh et al found that the age group most hit by occupational diseases was 50 years and above.^25 26^ The difference between our study, and those two is that their studies focused on former miners who were out of service, while we focused on in-service miners. In Zambia the law is that any prospective miners found with TB or have such history are not allowed to be hired, and those already miners, if found with TB are immediately removed from mine employment.^27^ This law automatically reduces the number of such patients in the working mine population.

### Occupational morbidity types

Wounds presented the highest proportion of the burden with a 30% incidence of all the cases in the period. This was followed by fractures, and amputations at 29%, and 17% respectively. In their study of compensation patterns in Zambia following occupational injuries, Siziya et al also found that wounds were the most common type of occupational injury.^19^ A similar study in Northwest Ethiopia found that cuts and fractures were prevalent at 24.6% and 2.5% respectively.^28^ Another study in India also found wounds as the leading injury type with 80.3% frequency. Fractures followed at a smaller frequency however of 7.7%.^12^ Our study findings are consistent with the results of the above studies.

In a much similar study also using workers compensation claim data from South Australia over a fifteen year period it was, nevertheless established that ligaments and tendon injuries were the leading injury type at 43.2%.^28^ Wounds, amputations, and all internal injuries were lumped up together at 22.1% as second highest. Although this study found ligaments and tendons as the leading type of occupational injury; wounds and fractures were still very prominent in the hierarchy. So their findings still concurred with ours.

### Industry

Our study found that manufacturing sector was carrying the highest burden at 27%. Mining, construction, and support services followed at 19%, 15%, and 13% respectively. This result is in agreement with the study findings in South Australia.^28^ Our study however is at variance with the findings of Namumba (2018) where the highest burden was found to be mining at 61.5% followed by construction 16.1%.^14^ The difference between the two could be that the other study was a local one only covering Kitwe district which is predominantly a mining town, and with a smaller sample size; whereas ours was a national study covering the whole country over a longer period of time. ILO(2012) in their country profile report on 2012 safety and health also showed different results from ours where mining was the highest followed by agriculture.^10^ Here the variance could be arising out of different industrial sector classification systems. Our classification of employers was based on United Nations International Standard Industrial Classification of All Economic Activities (ISIC);^29^ whereas the ILO report was based on the 2003 to 2007 summary reports of the Workers Compensation Fund Control Board employer classification system. Mining by nature is a more risky undertaking. It employs a lot of people exposed to a hazardous environment and machinery. Because of the potential danger this industry holds, in Zambia it is highly controlled and regulated. It has traditionally benefited from exposure to international occupational safety standards through the dominance of international mining conglomerates who are the primary investors in the sector. Thus, within each mining enterprise are highly skilled health and safety professionals providing controls and hence helping reduce accidents. Government regulatory departments (Mine Safety Department and Occupational Health & Safety Institute) also provide an overall inspection and tertiary supervisory role in ensuring safety of mining activities.

The manufacturing sector however, is relatively younger in the country with less supervision and controls, and by far less exposure to international occupational safety standards. It is dominated by small and medium enterprises, the majority of them family-owned, with little or no appetite to apply international occupational safety standards due to related compliance costs. Safety professionals are barely existent; and external controls are weak. This phenomenon could be one of the leading reasons why manufacturing industry in Zambia tends to be more occupationally hazardous than mining and others sectors.

### Region

In terms of the distribution of the burden of occupational morbidity by region, our study established that Lusaka region had the highest at 49%. This was followed by Copperbelt and Southern provinces at 34% and 6% respectively. This is consistent with the findings of CSO (2018) in their 2015/16 Zambia SAVVY report where they found that Lusaka province registered the highest number of all cases of mortality at 17.8%, followed by Copperbelt at 13.6%.^4^ In the same study, they also established that mortalities due to accidents and injuries alone were the highest in Lusaka at 20.9%, followed by Copperbelt at 19.7%. This national occupational hazard profile reflects the distribution of the occupational-safety-weakness of the manufacturing sector across the regions in the country.

## CONCLUSION

Our study went out to utilize workers compensation claims data as a proxy to understanding the patterns of occupational morbidity in Zambia over the period 2008 to 2018 by describing its major characteristics. Males in the age group 25 to 35 years were the most affected. The most prominent morbidity types affecting the workers in Zambia were wounds, fractures and amputations.

We established that the biggest contributor to the problem of occupational morbidities is the manufacturing sector. Poor regulation and controls of the manufacturing sector continues being the main driver of the problem. In terms of regional distribution Lusaka province which does not have mines but have the highest proportion of manufacturing sector leads in the burden of occupational morbidities than any other province in the country. The manufacturing sector should be subjected to stronger government regulation and inspection, with emphasis on compliance to relevant international occupational health and safety protocols.

This study did not cover morbidities from the informal sector. And we are not aware of any studies on the size of occupational morbidities from the informal sector. That said the challenge of occupational morbidities in Zambia must be bigger than what is presented in our findings. Future research should therefore include the magnitude of morbidities in the informal sector. Also it should investigate the quality of life of victims of occupational injuries and diseases.

## Supporting information

checklist

## Data Availability

Data set not available online. However, it may be made available to third parties upon reasonable request to the corresponding author.

## ETHICS APPROVAL AND DISSEMINATION

Ethical approval was granted by the University of Lusaka, medical Ethics Committee; and the Workers Compensation Fund Control Board Ethics Committee. This included a data extraction tool and data management plan to ensure adherence to general data Protection and health research regulations. The end study results will be published in an open access peer-reviewed journal.

## AUTHOR CONTRIBUTIONS

MZ

- Work conception. Data acquisition, analysis and interpretation.
- Manuscript drafting.
- Approval of final manuscript.
- Accountable for all aspects of the work regarding its accuracy or integrity.

PCB

- Data acquisition and interpretation.
- Critical revisions of manuscript for intellectual content.
- Approval of final manuscript.
- Accountable for all aspects of the work regarding its accuracy or integrity.

RM

- Conception and design of the work. Data analysis and interpretation.
- Manuscript drafting and critical revisions for intellectual content.
- Approval of final manuscript.
- Accountable for all aspects of the work regarding its accuracy or integrity.

JG

- Data interpretation
- Manuscript drafting and critical revisions for intellectual content.
- Approval of final manuscript.
- Accountable for all aspects of the work regarding its accuracy or integrity.

## ACKNOWLEDGEMENTS

This article is a part of the masters thesis submitted to the University of Lusaka in partial fulfillment of the requirements for the degree of Master of Public Health.

We gratefully acknowledge the management of the Workers Compensation Fund Control Board for approving us to use their compensation claims data for our research.

